# A rapid ONT-based sequencing approach to capture complete Ataxia-Mutomes (AtaxiaMutSeq)

**DOI:** 10.64898/2025.12.18.25342589

**Authors:** Tiyasha De, Mohammed Faruq

## Abstract

Hereditary ataxias are complicated neurological disorders with enormous genetic heterogeneity as well as the diverse genetic mechanism. Among different genetic mechanism, tandem nucleotide repeat expansion (TNRex) are the most common cause for genetic ataxias followed by single nucleotide variations in over 200 genes. The detection and the diagnosis of tandem nucleotide repeats in clinics and laboratories has been at large common in comparison with SNVs owing to the large number of the mutations in the respective genes they are found. The widely used platforms for detection of these mutations are capillary electrophoresis and Next generation sequencing based targeted gene panel or clinical or whole exome sequencing. Long read sequencers have been proven useful for detection of tandem nucleotide repeat expansions. We have evolved a method to detect in one experiment and on single platform the detection of TNRex and SNVs on Oxford Nanopr**e** Technology using adaptive sequencing approach. We were able to optimize the target region sequencing of both TNR loci and SNV-loci and validate the capture of both by detection of *FXN*-GAA repeats and pathogenic SNVs in *SETX*

## Introduction

Hereditary Ataxias (HAs) are neurodegenerative disorders primarily characterized by the degeneration of the cerebellum and its associated connections. Clinically, these disorders are predominantly marked by ataxia-related symptoms, including gait abnormalities, dysarthria, nystagmus, tremors, and upper limb incoordination. However, patients also present with a range of non-cerebellar symptoms1. The clinical manifestations of ataxia exhibit considerable heterogeneity and can be further complicated by co-occurring conditions such as hereditary spastic paraplegia (HSP), epilepsy, and hypo- or hyperkinetic movement disorders, which contribute to atypical ataxia phenotypes2. The overall prevalence of cerebellar ataxias is estimated at 1.5–4.0 × 10□ □ for spinocerebellar ataxias (SCAs)3 and 3.3 × 10□ □ for recessive ataxias^4^.

From a genetic standpoint, a significant portion of the hereditary ataxia diagnoses is attributed to tandem repeat expansion mutations (CNG), which account for approximately 30–40% of cases^5^, while another 30% are explained by conventional mutations^6^. Next-generation sequencing (NGS) has emerged as a critical tool for uncovering the genetic underpinnings of ataxias. Various sequencing approaches, including clinical exome sequencing, targeted panel testing, and whole exome sequencing, have proven invaluable for genetic diagnosis in previously uncharacterized cohorts, as well as for the discovery of novel genes associated with these disorders^7, 8^.

The classical approach to dissect genetic defect in ataxias are first to screen tandem nucleotide repeats expansions, like SCA1, SCA2, SCA3, SCA6, SCA7, SCA8, SCA12, SCA12, SCA10, SCA31, SCA36 and FRDA, and then to pursue the gene hunt using targeted gene panel sequencing or whole exome sequencing to screen 400-500 genes known to cause ataxia primarily (∼100 genes) or rarely with rest of the genes. In very rare scenario, CNV or structural changes have also reported as a causal mechanism. To investigate each cause of ataxia require a step-by-step or serial approach of testing by genetic methods like PCR, fragment analysis, Sanger sequencing, NGS, southern blotting etc. Short read sequencing may capture SNV and loci with smaller STR regions but will not detect the longer repeat expansions like FRDA etc. Likewise, Whole genome long-read sequencing is efficient in capturing repetitive DNA regions; however, a high error rate in sequencing bases may be undesirable outcome. However, given the technology advantage of Oxoford Nanopre technology platform offers a Third Generation Sequencing (TGS) approach by adaptive sequencing a one one-stop solution for capturing both repetitive region of SCA mutations as well SNV of several hundred genes in a cost effective and less data-intensive approach. There are very few reports of utilization of adaptive sequencing to capture both STR^9^, SNVs, CNVs and methylations using ONT platform.

We have performed adaptive sequencing for capturing nearly complete genetic spectrum of ataxia phenotypes on ONT. Our approach was successful in efficiently sequencing ∼70 STR regions, 6 full length gene sequencing of common ataxia causing genes by SNVs and 560 genes causing other rare forms of ataxias. Our approach exemplified a method that can be deployed in neurology clinic and has potential to diagnose a ataxia patient within 24-72 h hours in a cost effective manner.

## Methodology

### Sample recruitment

The pilot study was conducted using a simple control sample. The validation phase included two patient samples with prior molecular diagnosis of Friedreich’s Ataxia (FRDA) repeat expansion and Ataxia with Oculomotor Apraxia type 2 (AOA2), respectively. All individuals involved in this research study gave their informed consent to participate, and ethical approval was obtained from the Institutional Human Ethics Committee of CSIR-IGIB. Genomic DNA was extracted from peripheral venous blood samples using the salting-out technique (9).

### Library preparation

For each sample, genomic DNA (gDNA) at a concentration of 150 ng/μL was prepared in 20 μL of nuclease-free water (NFW). The gDNA was mechanically sheared by repeated passage through a 31-gauge insulin syringe to obtain fragments ranging between 8 and 15 kb. The fragmented DNA was subsequently processed for library preparation using the SQK-NBD114.24 kit. Although the SQK-LSK114 kit can be used as an alternative, the protocol was optimised with NBD114 to enable multiplexing. Several steps of the manufacturer’s protocol were modified to accommodate high DNA input. Specifically, the end preparation and DNA repair reaction volume was stoichiometrically increased to 25 μL, with an extended incubation time of 15 minutes at 20°C. Additionally, incubation time during native barcoding and adapter ligation steps was extended to 30 minutes. Libraries prepared using this optimised protocol were used for sequencing the samples. A total of 450 ng of each library was loaded onto individual R10 flowcells and sequenced with periodic reloading for up to 72 hours.

### Designing the bedfile for Adaptive Sequencing in Nanopore

The pilot study was designed to investigate 77 short tandem repeat (STR) loci, comprising 59 previously reported repeat expansion loci (10,11) and 19 novel candidate loci (12) identified through previous laboratory analyses. For each locus, the repeat region of interest was defined in the browser extensible data (BED) file with a (±) 5 kb flanking sequence. The total targeted genomic region in the pilot study encompassed 0.85 Mb.

The validation study employed a larger BED file that included genes known to cause ataxia through single-nucleotide variations (SNVs) or insertion-deletions (indels), Copy number variations (CNVs), and Structural Variations (SVs). This comprehensive design incorporated the entire genomic region of six common ataxia-associated genes (*SETX, ATM, SACS, SYNE1, SPG7*, and *SPG11*), along with the exonic regions of 253 genes, comprising 164 established ataxia-causing genes, 54 childhood-onset ataxia genes, and 35 candidate genes implicated in related disorders. These 35 candidate genes were selected based on prior whole-exome sequencing (WES) data from approximately 1,000 ataxia patients, in which recurrent pathogenic variants were observed in the cohort. In addition to these genes, the original set of 77 STR loci was retained in the BED file. Overall, the BED file comprised 3,537 genomic regions, of which 3,458 exonic regions spanned the 253 ataxia-associated and candidate genes. The total target genomic region in the validation study was 37.1 Mb. For each gene, exonic boundaries were defined using the longest transcript according to GENCODE (13), including the six entire gene regions with a (±) 5 kb flanking region.

### Nanopore Sequencing data analysis

FASTQ files generated in the “fastq_pass” directory were aligned to the hg38 reference genome using the Epi2Me Alignment workflow. Basecalling was performed using the dna_r10.4.1_e8.2_400bps_hac@v5.0.0 model. The resulting binary alignment/map (bam) files were subsequently analysed using the Epi2Me Human Variations workflow to detect SNVs and CNVs. The resulting variant call format (vcf) files were annotated using ANNOVAR (14) and further analysed to identify pathogenic sequence variants. We further used Straglr to estimate repeat lengths at each STR locus. While Straglr (15) employs a default BED file comprising 38 known pathogenic loci, a custom tandem repeat BED file with the chromosomal locations and repeat motifs was provided to Straglr to enable repeat length estimation across all 77 loci of interest. The resulting tab-separated values (tsv) file was analysed for repeat expansions.

## Results and Discussion

### Pilot study

After 72 hours of sequencing with 2 subsequent loadings of the library, the total data generated was approximately 13Gb with a N_50_ of 7.17 kb. The mean coverage of the sample across the bedfile was roughly 15x. The reads had a mean read quality of 13.7, a mean read length of 1,799.8 kb and a maximum read length of 166,207 kb. The FASTQ pass files were taken and aligned to hg38, and the number of repeats in each locus was estimated using Straglr. The distribution of repeats in each locus for the specified repeat motif can be visualised in Figure 1(A). Of the 77 STR loci under investigation, Straglr couldn’t estimate the number of repeats in 14 loci in this sample.

**Figure 1:**
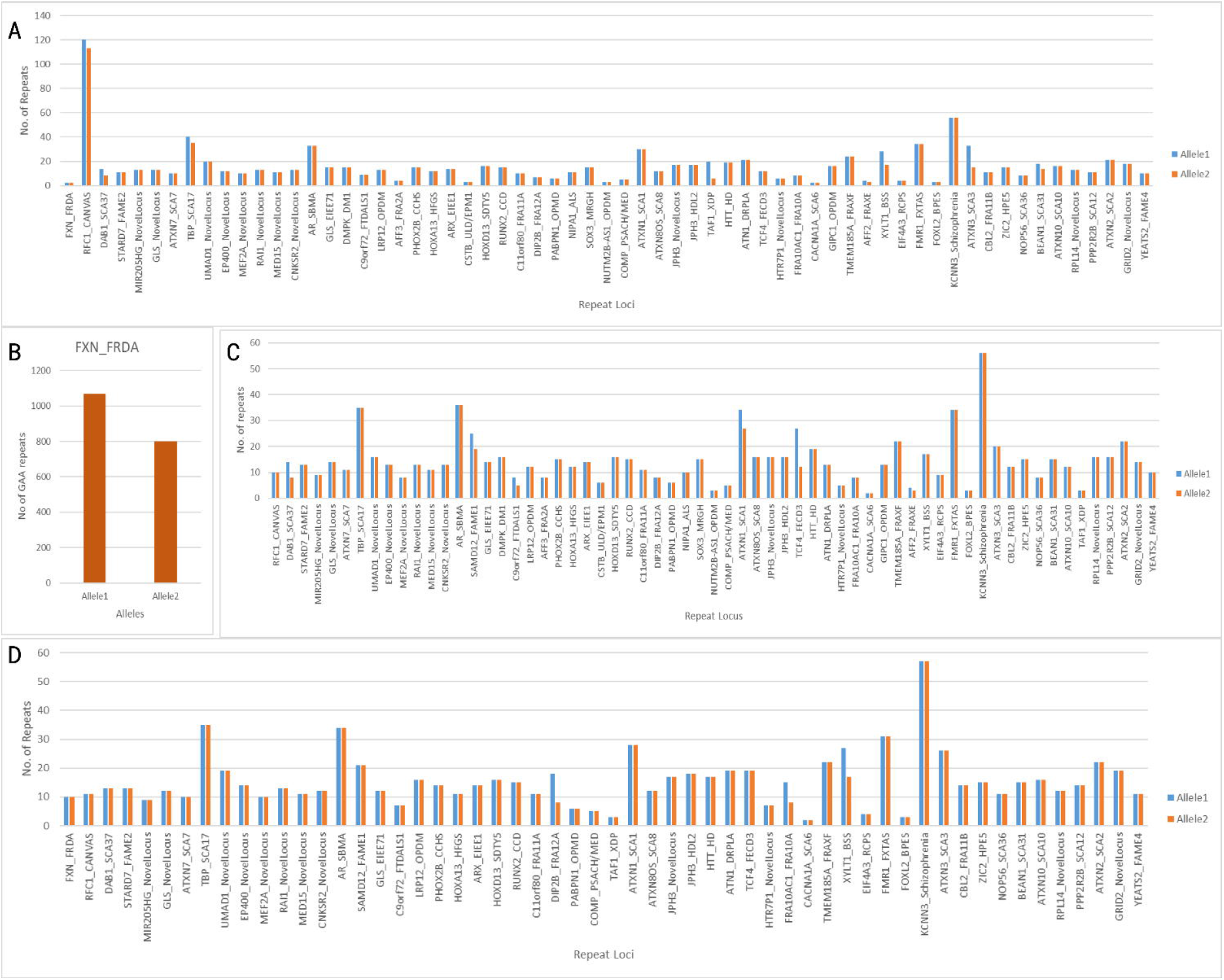
The figure represents the distribution of repeats in each of the 77 STR disease-causing loci for the specified repeat motif. Repeats estimated in allele 1 and allele 2 are represented, respectively in each pathogenic disease-causing locus in the X-axis while the Y-axis depicts the repeat numbers; (A) Represents the repeat distribution across 63 loci estimated using Straglr in the control sample in the pilot study; (B) Represents the repeat distribution in the pathogenic FXN gene locus in a patient diagnosed with FRDA from the validation cohort; (C) Represents the repeat distribution across 63 other loci estimated using Straglr in the FRDA patient sample in the validation cohort; (D) Represents the repeat distribution across 56 loci estimated using Straglr in the AOA2 patient sample in the validation cohort.

### Validation study

The validation cohort comprised patient samples previously confirmed to harbour FRDA-GAA repeat expansion and pathogenic point mutation associated with AOA2. This validation cohort was used to evaluate the efficiency of Nanopore adaptive sequencing as a cost-efficient approach for simultaneous detection of repeat expansions and pathogenic mutations in ataxia patients. The FRDA patient sample was specifically included because the disorder is caused by a GAA repeat expansion in intron 1 of the *FXN* gene, with pathogenic alleles harbouring repeats over 1,000 repeating units. This wide spectrum of repeat lengths, alongside its autosomal recessive mode of inheritance, made FRDA an ideal model to assess the efficiency of adaptive sequencing to accurately identify large and complex repeat expansions of this magnitude.

### FRDA positive patient sample

After 72 hours of sequencing with 2 subsequent loadings of the library, the total data generated was approximately 10Gb. The mean coverage of the sample across all 77 repeat expansion loci was roughly 28x, while the average coverage across all the genes with ataxia-causing point mutations was 20x. Of the 77 STR loci under investigation, Straglr couldn’t estimate the number of repeats in 13 loci in this sample. The tsv output file from Straglr validated the biallelic repeat expansion in intron 1 of the *FXN* gene with 800 and 1068 GAA repeats in allele 1 and allele 2, respectively. The distribution of repeats in FRDA causing *FXN* gene is depicted in can Figure 1(B) while the repeat distribution across all the other loci for the specified repeat motif can be visualised in Figure 1(C). During SNV-indel analysis, a total of 20,457 variants were identified, with 16,258 SNVs and 4,221 Indels. The number of SNVs and indels is computed independently, and their sum could be different from the number of records due to multiallelic variants. No pathogenic variants were identified in the patient sample. In the SV analysis, a total of 79 insertions, 59 deletions, and 6 other types of SVs (inversions, duplications, and translocations) were identified, with most occurrences in chromosome 20. Figure 2(A) depicts the occurrence of these SVs across all the chromosomes.

**Figure 2:**
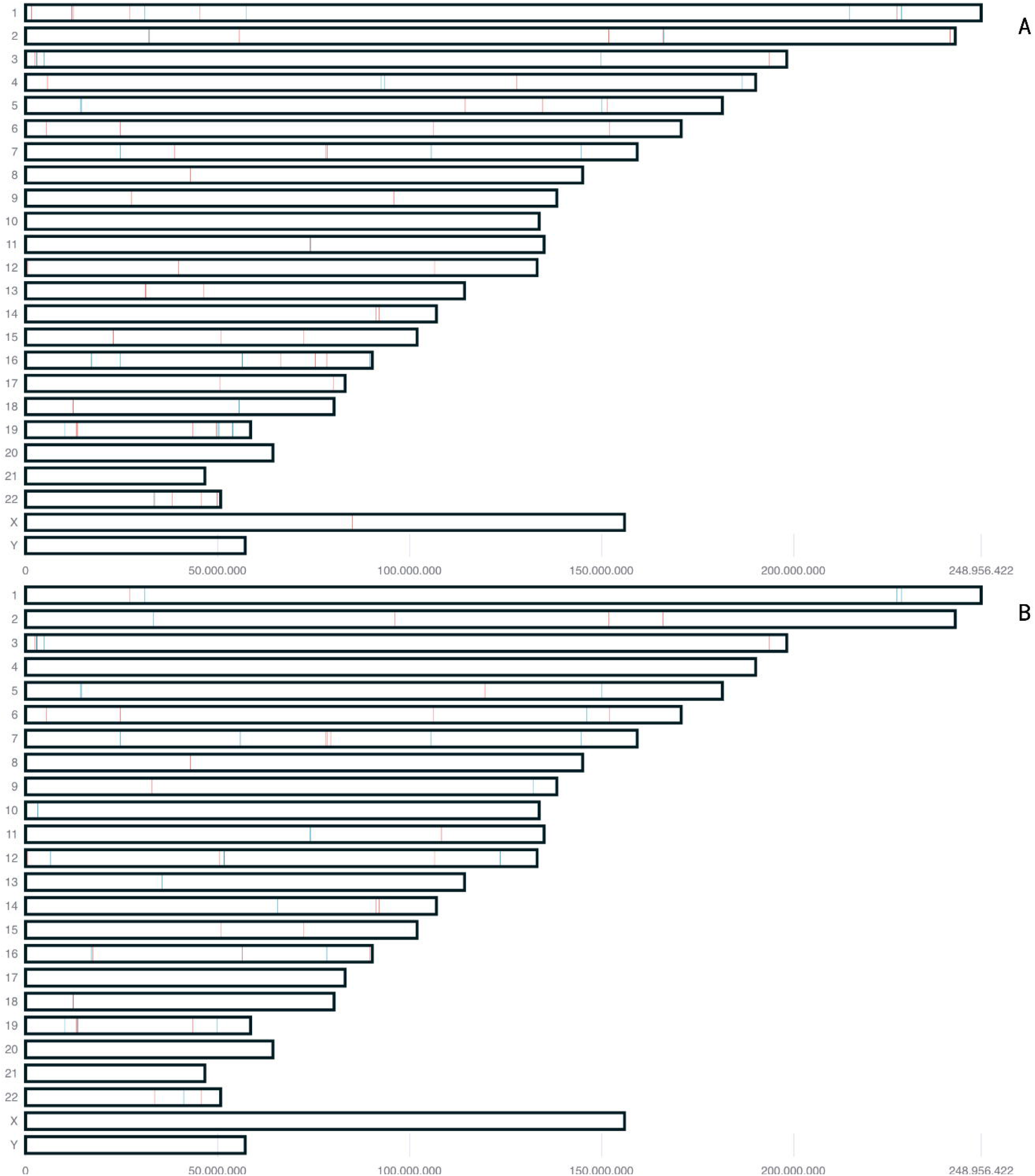
Karyogram representing the chromosomal hotspots of structural variants, both insertions (red) and deletions (blue), across all the chromosomes in the genome. (A) Represents karyogram of FRDA patient sample; (B) represents the karyogram of AOA2 patient sample.

### AOA2 patient sample

After 72 hours of sequencing with 2 subsequent loadings of the library, the total data generated was approximately 6.5Gb. The mean coverage of the sample across the BED file was roughly 8x. There are no pathogenic repeats identified in the patient. The distribution of repeats in each locus for the specified repeat motif can be visualised in Figure 1(C). Of the 77 STR loci under investigation, Straglr couldn’t estimate the number of repeats in 21 loci in this sample. A total of 24,018 variants were identified, with 19,005 SNVs and 5,019 Indels. The number of SNVs and indels is computed independently, and their sum could be different from the number of records due to multiallelic variants. We validated the previously identified pathogenic frameshift deletion, p.Lys796fs, at genomic position chr9:132,329,207 (hg38). In the SV analysis, a total of 40 insertions, 37 deletions, and 22 other types of SVs (inversions, duplications, and translocations) were identified, with most occurrences in chromosome 19. Figure 2(B) depicts the occurrence of these SVs across all the chromosomes.

## Conclusion

Among the 77 STR loci under investigation, 13 loci were consistently observed across both the pilot and validation cohorts for which repeat length estimation could not be reliably obtained. These loci need further investigation for such an unvaried pattern in future experiments. The validation cohort demonstrates that Nanopore adaptive sequencing, in combination with the Epi2Me human variation workflow, is a robust and reliable approach for investigating genetically complex disorders like ataxia. This strategy enables a streamlined, single-nucleotide variant and structural variation analysis, facilitating comprehensive, end-to-end genetic diagnosis. Importantly, the strength of the protocol lies in its capacity for sample multiplexing using the SQK-NBD114 kit, which significantly enhances the cost-efficiency and enables high-throughput analysis of critical patients in both research and clinical diagnostic settings.

### Ethics statement

Informed consent was obtained from participating subjects. The study was approved by Institutes Human Ethics Committee (IHEC), CSIR-IGIB for GAP240 project.

## Acknowledgement

We sincerely acknowledge funding support from Indian Council of Medical Research (ICMR) funded project, 5/4-5/5/Ad-hoc/Neuro/220/NCD-I (GAP240) AtaxiaChanger project.

## Author Approval

all authors have seen and approved the manuscript.

## Competing Interests

None to report.

## Data Availability Statement

All data will be available on request from corresponding author

## Figure Legends

**Table 1:**
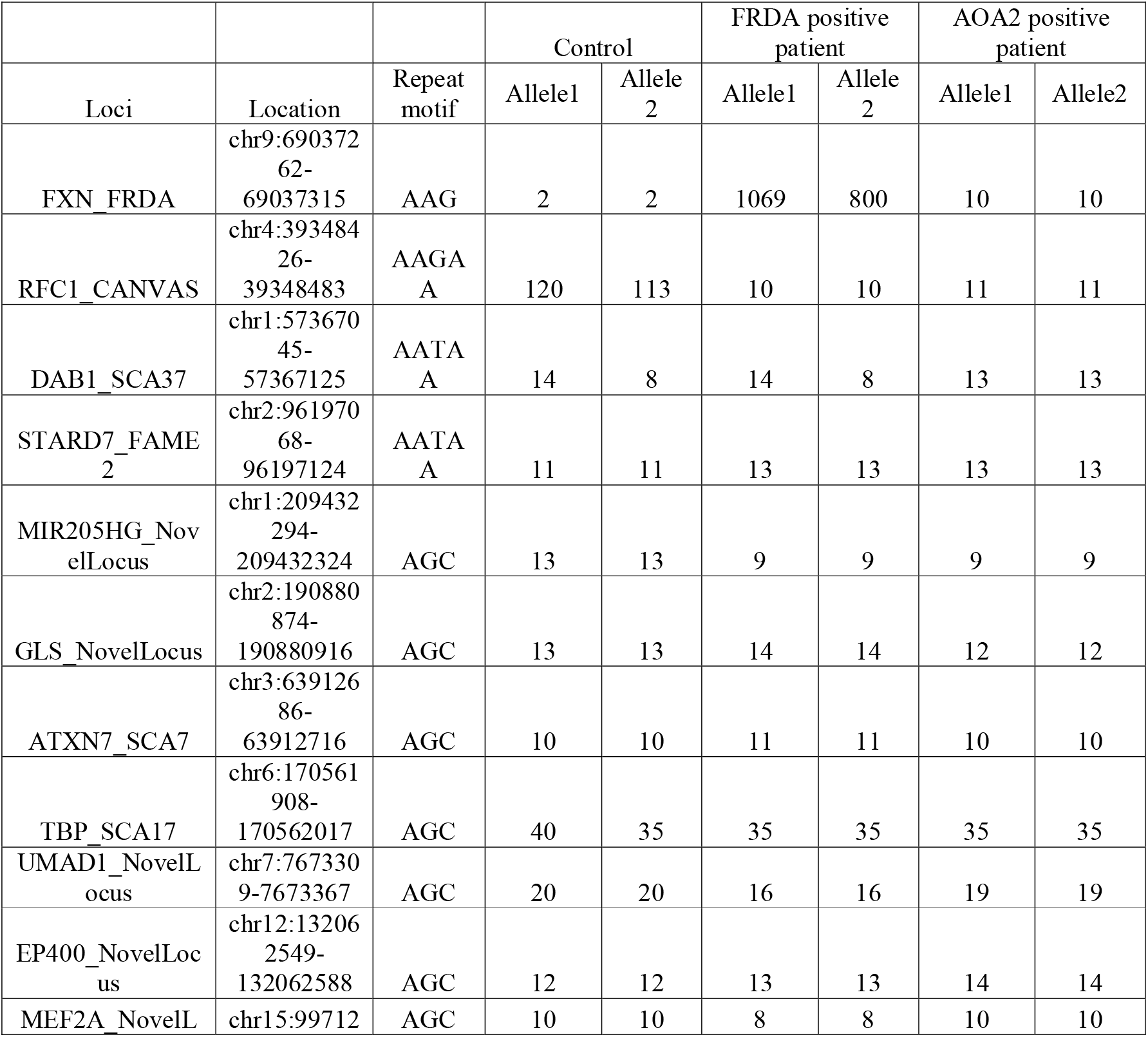
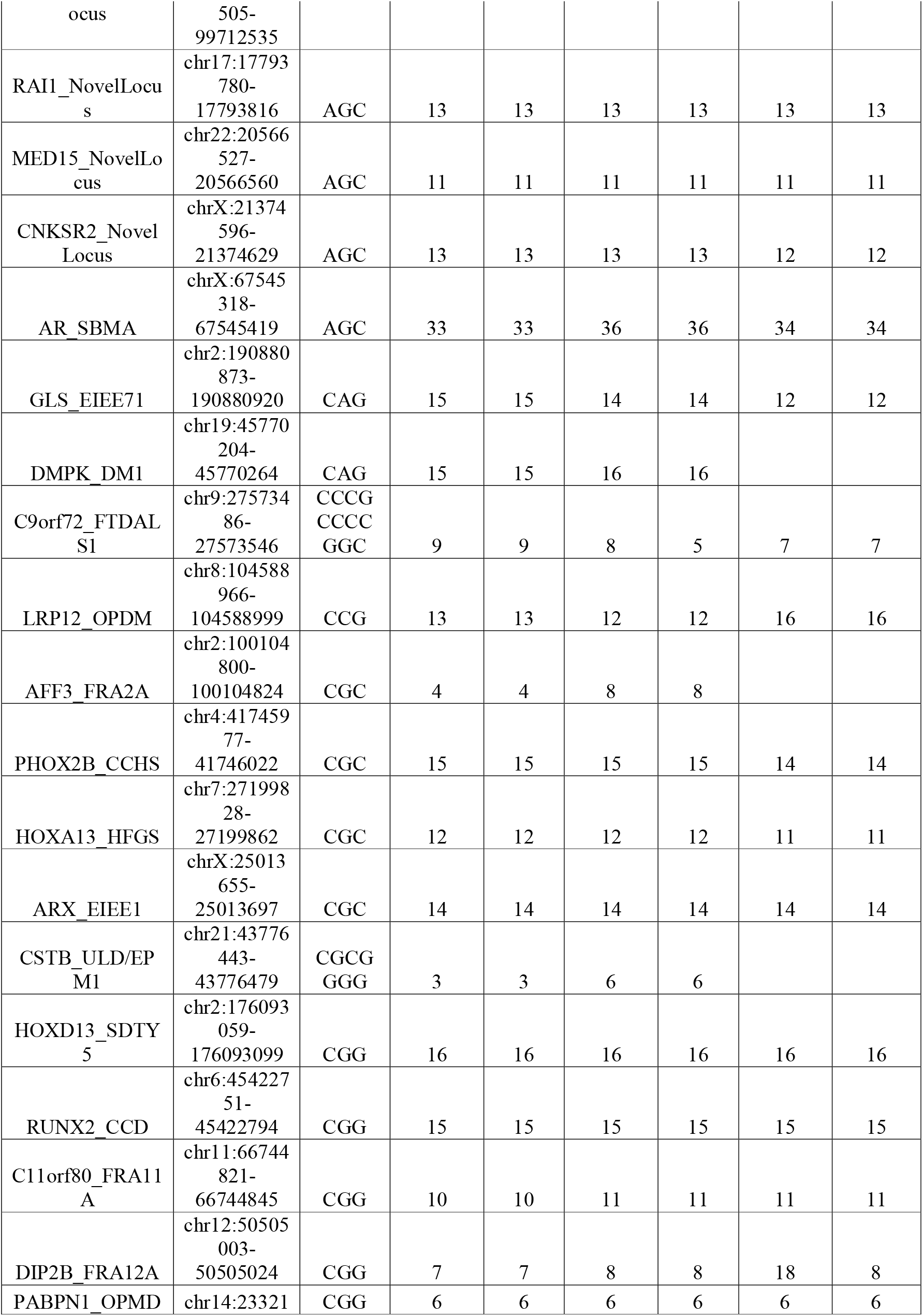
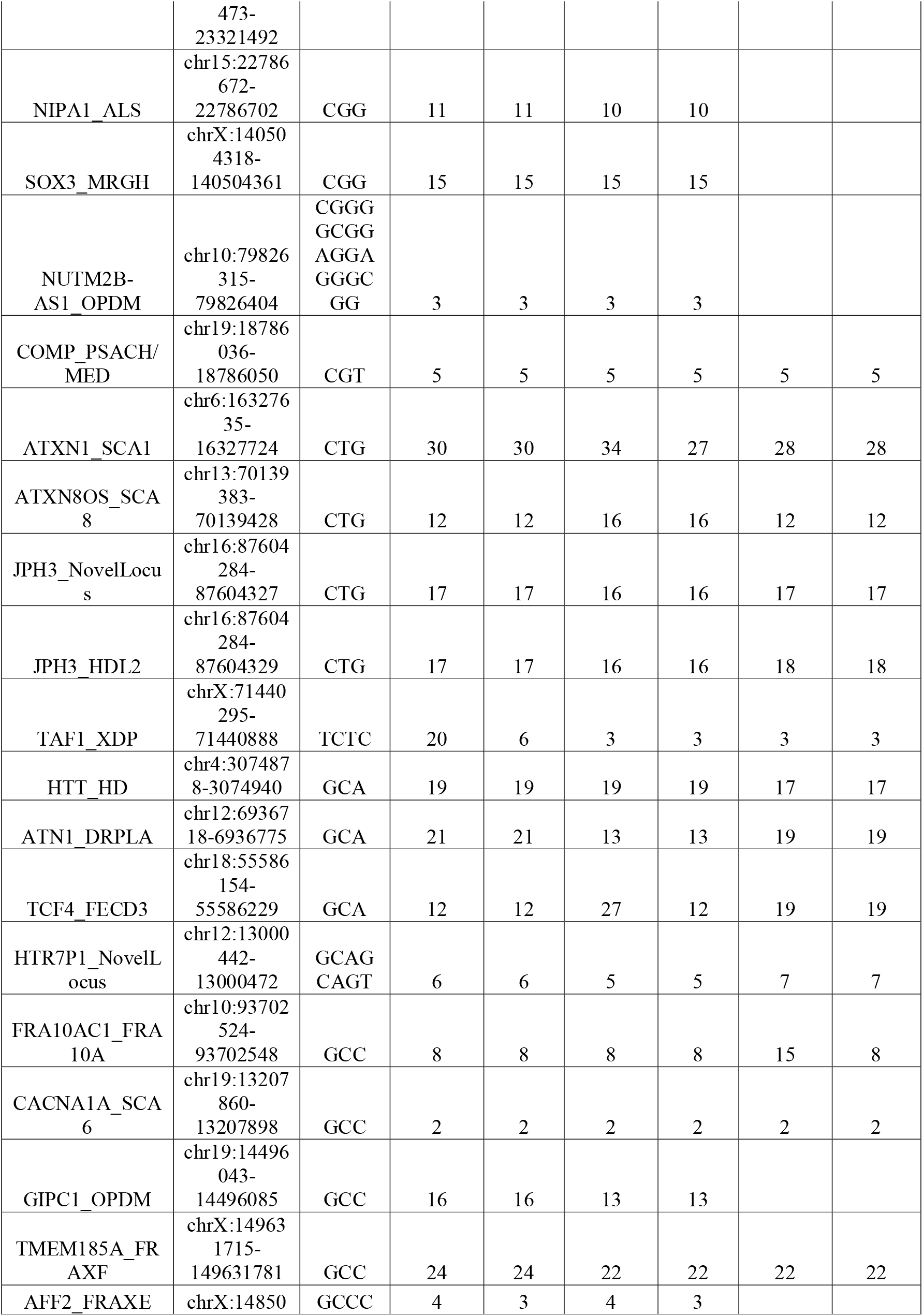
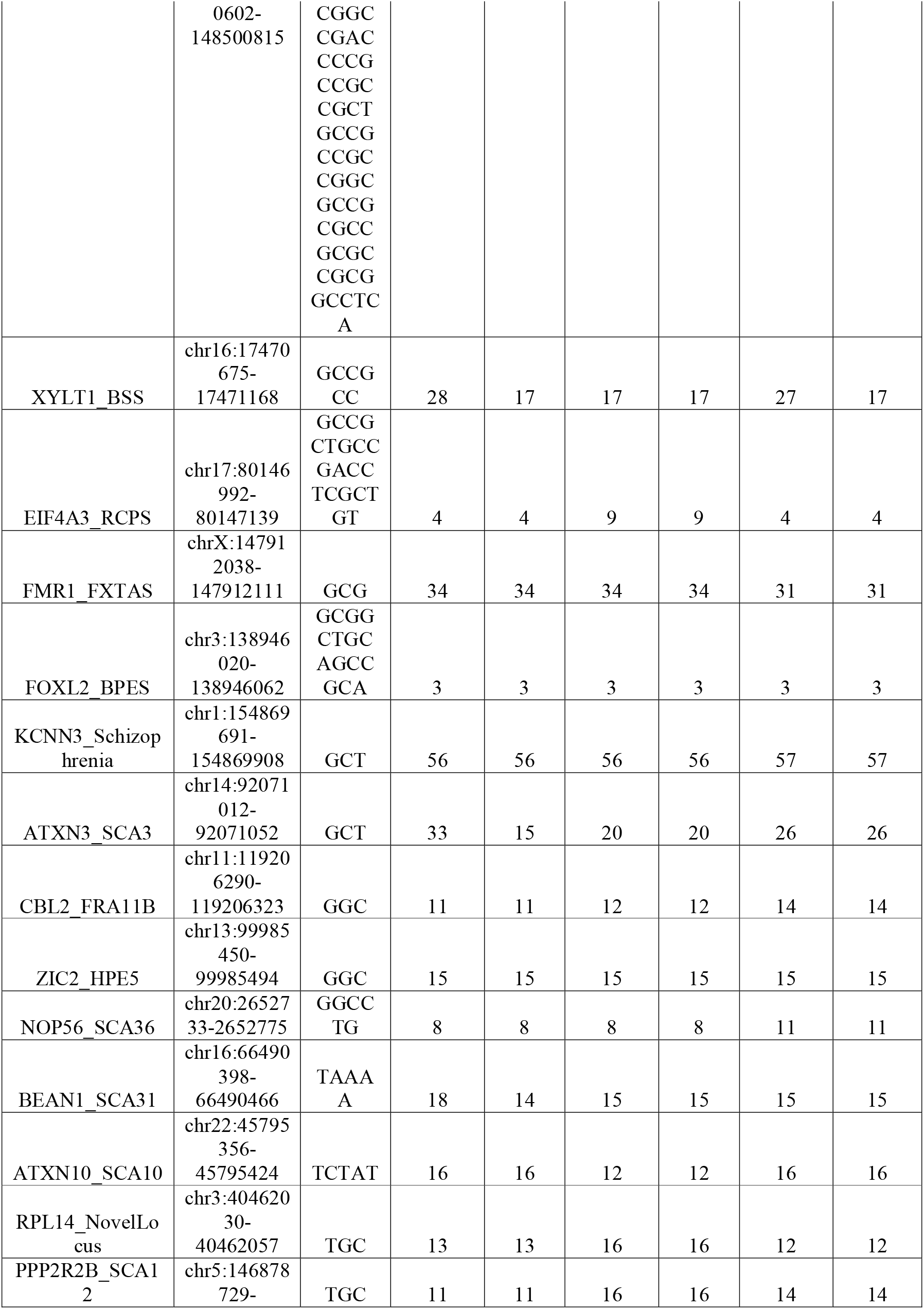
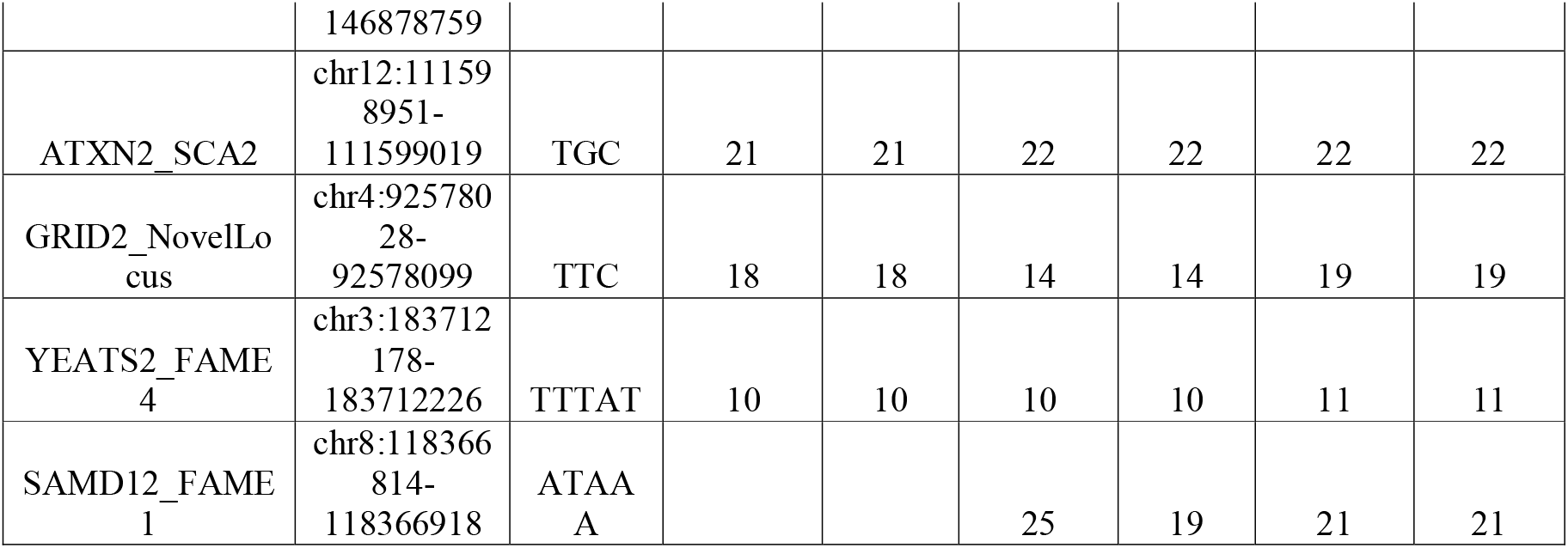
Shows the distribution of estimated repeats using Straglr across each locus for the given repeat motif.

## Notes

### Competing Interest Statement

The authors have declared no competing interest.

